# The Effect of Treating Hearing Loss with Hearing Aids on Plasma Biomarkers of Alzheimer’s Disease and Related Dementias

**DOI:** 10.1101/2025.11.19.25340558

**Authors:** Lachlan Cribb, Margarita Moreno Betancur, Julia Sarant, Rory Wolfe, Matthew Paul Pase, Gary Rance, Michelle M. Mielke, Anne M. Murray, Alice Owen, Robyn L Woods, Zhen Zhou, Zimu Wu, Kerry M Sheets, Trevor T.-J. Chong, Raj C Shah, Joanne Ryan

## Abstract

**Background:** Promising evidence indicates that treating hearing loss with hearing aids (HAs) could reduce dementia risk. We extend this evidence by investigating the effect of HAs on plasma biomarkers of Alzheimer’s disease and related dementias (ADRD).

**Methods:** We emulated two target trials using observational data from Australian participants of the ASPREE study. Eligible participants had self-reported hearing problems, no past HA use, and were dementia-free. HA prescriptions and frequency of HA use were measured by questionnaire. Phosphorylated-tau181 (pTau181), neurofilament light chain (NfL), glial fibrillary acidic protein (GFAP), and amyloid-β (Aβ) 42/40 were measured after approximately 6-8 years. We estimated the effect of new HA prescription (first target trial) and the frequency of HA use (second target trial) using targeted maximum likelihood estimation, with multiple imputation for missing data.

**Results:** Across imputed datasets, a median of 2842 eligible individuals were included (mean age 75 years, 48% female), with a median of 735 receiving a new HA prescription. Among survivors, the estimated mean differences comparing HA prescription and no HA prescription were 1.8 pg/mL (95% CI: -0.6, 4.1), 0.1 pg/mL (-7.8, 8.0), -2.2 pg/mL (-14.5, 10.1), and -0.7 (-2.6, 1.2) for the concentrations of pTau181, NfL, GFAP, and (Aβ42 x 1000)/Aβ40, respectively. Mean differences did not differ substantially across levels of potential baseline effect modifiers, including APOE-ε4 genotype and cognition.

**Conclusion:** In community-dwelling older people with hearing loss and no dementia, we found minimal effects of HA prescription and frequency of HA use on plasma ADRD biomarkers after a 7-year follow-up.

## INTRODUCTION

Treating hearing loss with hearing aids (HAs) is a promising strategy for limiting cognitive decline and preventing dementia.^1^ There are several potential mechanisms by which this benefit could occur, including sparing cognitive resources that would otherwise be diverted by the demands of listening in difficult environments, reducing the psychosocial sequelae of hearing loss (e.g., social withdrawal and depression), and limiting the interaction between the neurological effects of hearing loss and neuropathology.^2,3^

The ACHIEVE trial investigated an intervention consisting of HA use and accompanying technologies for cognitive change in older US adults with hearing loss. The trial found little overall effect of the intervention on cognitive change over 3 years.^4^ To complement this evidence, our recent study investigated the effect of HAs on cognitive change and dementia risk over a 7-year follow-up in cognitively healthy older adults.^5^ We estimated that 7-year dementia risk was 33% lower in those who received a new HA prescription by the start of follow-up, compared to those who did not, though differences in longitudinal cognitive change were modest and equivocal.^5^ In ACHIEVE and in our study, effects on cognitive outcomes tended to be greatest in those at high risk for cognitive decline.^5,6^

Phosphorylated-tau181 (pTau181), amyloid-β42/amyloid-β40 (Aβ42/Aβ40), neurofilament light chain (NfL), and glial fibrillary acidic protein (GFAP) have emerged as key biomarkers of Alzheimer’s disease and related dementias (ADRD). The plasma concetrations of these biomarkers are closely correlated with ADRD pathology^7^ and are prognostic for cognitive decline and dementia.^8,9^ Investigating these plasma biomarkers as outcomes allows for the assessment of treatment effects at an early disease stage (i.e., before clinical endpoints, like dementia, occur) and can provide insight into treatment mechanisms.^10^ Accordingly, we planned to extend our previous study into HA effects on cognition and dementia risk by investigating these biomarkers as outcomes.

The primary aim was to investigate the effect of HA prescription and the frequency of HA use on the mean concentration of plasma biomarkers of ADRD. Additionally, hypothesising that HA use would influence the development of *marked* (i.e., highly elevated) levels of neuropathology, we also aimed to estimate the effect of HA prescription and frequency of HA use on shifting the 90^th^ percentile (10^th^ percentile for Aβ42/Aβ40), rather than the mean, of the biomarker outcome distributions. Finally, we aimed to investigate effect modification by pre-treatment ADRD risk factors, including the baseline plasma biomarkers, baseline cognition, and APOE-ε4 genotype.

## METHODS

### Observational data source

We used observational data from Australian participants of the ASPREE study and the Australian Longitudinal Study of Older Persons (ALSOP). ASPREE was a randomised, placebo-controlled trial investigating low dose aspirin for disability-free survival in 19,114 older individuals in Australia and the United States.^11,12^ In Australia, participants were recruited between 2010 to 2014 aged ≥ 70 years through collaboration with their primary care physician. ASPREE participants were followed prospectively until the trial ended in 2017 and thereafter at annual visits as part of an observational extension (ASPREE-XT).^13,14^ ALSOP was a substudy in 14,892 Australian ASPREE participants that collected information on medical and social health factors, including hearing function and hearing loss treatment, by questionnaire.^15^ The first ALSOP questionnaire was completed near ASPREE baseline and the second was completed approximately three years later (**Figure 1**).

**Figure 1.**
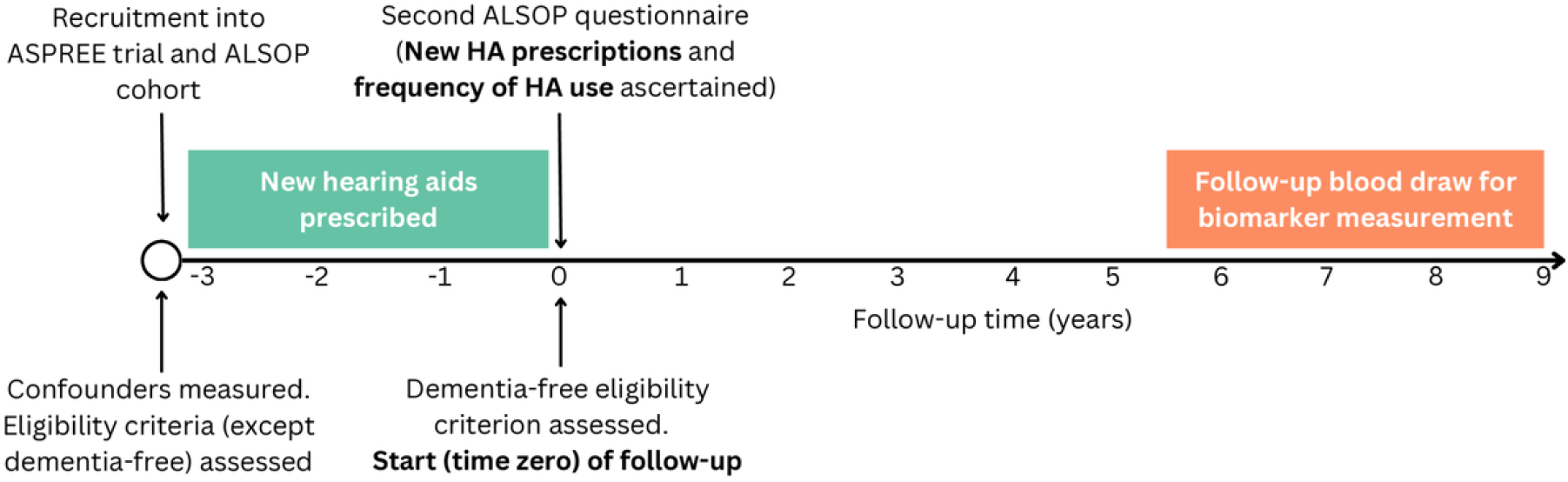
Study design for emulation of each target trial

Data on self-reported HA prescriptions and frequency of HA use were drawn from the ALSOP surveys. For context, HAs in Australia are provided free of charge to eligible pensioners (aged ≥67 years) and to disability healthcare card holders through the Hearing Services Program. Subsidised funding of HA is also provided by Australia’s Department of Veterans’ Affairs and private health insurers. HAs within this study period would have been prescribed by an audiologist within a hearing clinic as part of standard clinical practice.

In ASPREE participants, blood samples were drawn for the measurement of plasma biomarkers of ADRD at baseline (n=11,961) and again approximately 10 years later (n=5,243). The protocol for sample collection, processing, and storage has been described in detail.^16,17^ All biomarker assays were completed at the Advanced Research and Diagnostic Laboratory (ARDL) at the University of Minnesota, USA, on the Quanterix HD-X platform in 2023. The concentrations of Aβ40, Aβ42, NfL, and GFAP were measured on the Simoa® Human Neurology 4-Plex E (N4PE) platform. pTau181 was measured using the Simoa® pTau181 v2 assay. Inter-assay and intra-assay coefficients of variation from a subsample of duplicated measurements were ≤5% and <7% for all biomarkers, respectively.^17^

Finally, we also utilised data from a subset of ASPREE and ALSOP participants (n=1,166) that underwent baseline audiometry assessments as part of the ASPREE-Hearing substudy.^18^ The use of this data for the primary and sensitivity analyses is described below. ASPREE and ASPREE-XT were approved by the ethics review board at each participating institution. All participants provided written informed consent.

### Protocols for the target trials and their observational emulation

This observational analysis is designed to emulate two target trials (i.e., the hypothetical randomised trials that would be performed to answer our research question)^19–21^ with different treatment strategies. The protocols for these target trials and their emulation using observational data are described in **Table 1**. The overall design of the observational emulation is summarised in **Figure 1**.

**Table 1:** Protocol for the target trials and their observational emulation.

| *Protocol component* | *Target trials* | *Observational emulation* |
| --- | --- | --- |
| *Eligibility criteria* | <u>Inclusion criteria:</u><br>• Aged 70+ years<br>• Moderate or more severe hearing loss (better-ear 4-frequency pure tone average ≥ 30 dB HL)<br>• 3MS examination score $> 77$<br>• Community dwelling<br><u>Exclusion criteria:</u><br>• Dementia<br>• Inability/severe difficulty or requiring assistance in performing any activity of daily living<br>• Past hearing aid (HA) or cochlear implant<br>• Contraindication for HAs<br>• Unwillingness to wear HAs | Same as target trial, except<br>• Self-reported hearing problems is used instead of moderate or more severe hearing loss<br>• Consent for participation in ASPREE and ALSOP is required<br>• No exclusion for HA contraindications or unwillingness to wear HAs<br>• Additional ASPREE study exclusion criteria<sup>1</sup><br>Eligibility was assessed using variables measured at entry into the ASPREE/ALSOP cohort, except for remaining dementia-free which were assessed at time zero (defined as 3 years later; Figure 1). |
| *Treatment strategies* | *Target trial 1*<br>1. Use HAs<br>2. Do not use HAs<br>*Target trial 2* | Same as target trial. |
|  | 1. Never use HAs<br>2. Use HAs rarely/sometimes (>0 and <4 times per month)<br>3. Use HAs often/always ($\geq 4$ times per month) |  |
| *Assignment procedure* | Participants are randomly allocated to one of the treatment strategies without blinding. | We classified participants to the treatment groups consistent with their hearing aid prescription/frequency of use data, self-reported in the second ALSOP questionnaire (Figure 1) |
| *Outcomes* | Plasma concentrations of pTau181, NfL, GFAP and Aβ42/Aβ40 | Same as target trial. |
| *Follow-up* | *Starts (time zero):* At the time of determining eligibility criteria and treatment assignment. | *Starts (time zero):* At the 3<sup>rd</sup> year of the ASPREE study (when the second ALSOP questionnaire is completed, and new HA prescriptions are identified). |
|  | *Ends:* At follow-up blood draw (after 7 years on average), loss to follow-up, or death. | *Ends:* At follow-up blood draw (after 7 years on average, i.e. 10 years after recruitment into ASPREE/ALSOP), loss to follow-up, or death. See Figure 1. |
| *Causal contrast* | Intention-to-treat effect in those who survive until biomarker measurement. | Observational analogue of the intention-to-treat effects. In the emulation of the first target trial, this is the effect of HA prescription. In the emulation of the second target trial, it is the effect of initiating a given frequency of HA use. |

3MS = Modified Mini-Mental State examination. <sup>1</sup> Including previous cardiovascular disease events and not being expected to survive for the next 5 years (ASPREE exclusion criteria are described in the primary trial report<sup>6</sup>). See main text for identifying assumptions and statistical analysis.

#### Eligibility criteria

In the target trials, eligible participants are those who are dementia free, have significant hearing loss (better-ear 4-frequency [0.5–4 kHz] pure tone average [PTA] of ≥30 dB HL), no past HA use, and do not have contraindications to HAs (e.g., profound/total hearing loss).

The emulation applied the same criteria, with two exceptions. First, as audiometric data were only available for a small proportion of the sample, we used presence of self-reported hearing problems to emulate significant hearing loss. In the subset reporting hearing problems and with audiometry data available, the median hearing loss was 29 dB HL. Most (81%) had ≥20 dB HL. Second, those with contraindications to HAs were not excluded, as these data were unavailable. Participants were assessed for the hearing-related eligibility criteria at ASPREE/ALSOP baseline and were required to be dementia-free at the start (time zero) of follow-up of the emulation, defined as the third year of the ASPREE study (**Figure 1**).

#### Treatment strategies

For the first target trial, the treatment strategies are **use HAs** (at any frequency) and **do not use HAs** throughout follow-up. For the second target trial, to explore the dose-response relationship between HA use and the outcomes, the treatment strategies are **never use HAs**, **use HAs rarely/sometimes (>0 and <4 times per month)**, and **use HAs often/always (**≥**4 times per month)** throughout follow-up. The strategies are the same in the emulation.

#### Assignment

In the target trials, participants are randomly allocated to one of the treatment strategies without blinding. In the emulation of the first target trial, participants were classified into the “use HAs” strategy if they reported a new HA prescription in the second ALSOP questionnaire (i.e. at the third year of the ASPREE study, defined as time zero per above; **Figure 1**) and to the “do not use HAs” strategy otherwise. In the emulation of the second target trial, participants were assigned to strategies based on their reported frequency of HA use in the second ALSOP questionnaire, with those reporting no HA prescription or no HA use assigned to the “never use HAs” strategy.

#### Follow-up

In the target trial, follow-up begins at treatment assignment and ends at blood collection for biomarker measurement (approximately 7 years later) or death, whichever occurs first. In the emulation, follow-up began at the 3^rd^ year of the ASPREE study, when new HA prescriptions were first reported, and ended in the same manner as the target trial (**Figure 1**). Loss to follow-up in the emulation was defined as missing outcome data not due to death.

#### Outcomes

Outcomes were the plasma concentrations of pTau181, Aβ42/Aβ40, NfL, and GFAP.

#### Causal contrasts

For the target trials, the primary causal contrast is the intention-to-treat (ITT) effect (i.e., the effect of treatment allocation, irrespective of adherence) on the mean difference scale in survivors. We chose to investigate this survival conditional effect, rather than an effect that is “unconditional” on survival, as the latter would require imputation for post-death outcomes, and this is not generally meaningful.^22^ Secondary contrasts of interest are the ITT effect of HAs on the quantile difference scale, specifically the difference in the 90^th^ percentile (or 10^th^ percentile in the case of Aβ42/Aβ40) of the biomarker distribution between strategies, conditional on survival.

For the emulation of the first target trial, the ITT effect is defined by using HA prescription to emulate allocation. For the second target trial, the ITT effect is defined by using initiation of a given frequency of HA use to emulate allocation. We refer to these as “observational analogues” of the ITT effect.^19^ All contrasts were conditional on participants being alive when the follow-up blood draws began.

### Identifying assumptions

We assumed that there was no residual confounding after adjusting for baseline covariates including factors associated with hearing aid initiation,^12,13^ self-rated hearing function, dementia risk factors,^1^ clinical factors, the pre-treatment plasma ADRD biomarkers, a biomarker-based dementia risk score (see below), and APOE-ε4 genotype. See **eTable 1** for all covariates. In all analyses, we additionally adjusted for audiometric hearing loss (the handling of missing data in this and other covariates is described below). Further identifying assumptions, concerning missing data, positivity, consistency, and measurement error, are described in the **eMethods**.

### Statistical analysis

Statistical analysis followed a prespecified plan available at https://osf.io/9faew/overview. Analysis was performed using R version 4.3.3.^23^ The analysis code is available at https://github.com/Lachlan-Cribb/Hearing-aids-biomarkers.

#### Biomarker-based dementia risk score

Using data from ASPREE study participants who were not eligible for inclusion in our analytic sample (n = 8,329), we fitted a model for the 10-year risk of dementia based on the baseline plasma biomarkers. Large estimated risk scores indicated high-risk baseline biomarker profiles. We used this risk score to assess for effect modification and to adjust for confounding by baseline preclinical disease. See **eMethods** for details.

#### Missing data

Missing data is summarised in **eTable 2**. There were missing data in exposures (18%), covariates (mostly <10%) and the eligibility criterion of self-reported hearing problems (5%). Audiometric data were missing for most participants (91%), principally due to calendar time as the ASPREE Hearing sub-study began during the latter stages of ASPREE recruitment.^18^ The follow-up plasma ADRD biomarkers were missing for 55% of surviving participants. This proportion of missing outcome data is expected given the approximately 10-year gap between ASPREE enrollment and the follow-up blood draw and consequently the age of participants at this point (mean of 85 years). The main reasons for missing outcome data were: i) participants were no longer interested in or able to attend in-person the ASPREE-XT visit when the follow-up blood draw occurred and ii) loss to follow-up.

We used multiple imputation (MI) to handle all missing data. We chose to use MI rather than restricting the analysis to those with complete data as i) MI allowed us to incorporate longitudinal auxiliary variables (and thereby to satisfy missing data assumptions; see **eMethods**) and ii) MI allowed us to adjust for audiometric hearing in the analyses, despite this only being available for a small proportion of the sample. MI was implemented using the predictive mean matching method in the R package *mice*.^17^ Pre-specified imputation models included outcomes, covariates, treatment, and a subset of longitudinal auxiliary variables (see **eTable 3**). To ensure compatibility with the main analysis method (described below), non-linear and two-way product terms for key variables were included.^24^ The subsets of auxiliary variables, non-linear and product terms (excluding those between treatment and pre-specified effect-modifiers, which were always included) to include in imputation models were determined using LASSO variable selection.^19^ In the procedure, quadratic terms and product terms were passively imputed.^18^ MI was performed once to impute missing data for all analyses (primary and sensitivity).

We used bootstrapping (250 samples) followed by multiple imputation to obtain confidence intervals.^20^ Two imputed datasets were created within each bootstrap sample. Point estimates and confidence interval limits were computed by pooling across imputed datasets and bootstrap samples using established formulas.^20^

#### Estimation

Causal effects on the mean difference scale were estimated using targeted maximum likelihood estimation (TMLE).^25^ We used pre-specified generalised linear models for estimating the conditional mean of the outcome and the propensity score (see **eTable 1**). Due to non-convergence in some bootstrap samples, product terms that were pre-specified for inclusion in the treatment model (e.g., between sex and hearing variables) were removed. Propensity scores were truncated at sqrt(n * ln(n)) / 5, where n is the sample size.^26^ To estimate effects on the quantile difference scale, we used an inverse probability weighted estimator.^27,28^

To investigate modification of the effect of HA prescription (on the mean difference scale), we used TMLE to estimate the parameters of a marginal structural model.^29^ The marginal structural model included treatment, the effect modifier (modelled with restricted cubic splines with knots at the 10th, 50th, and 90th percentiles for continuous effect modifiers), and their product(s). Mean biomarker outcomes under each treatment strategy were estimated from this marginal structural model across levels of the effect modifier.

### Sensitivity analyses

We conducted sensitivity analyses to examine underpinning assumptions:

1. We applied the same 30dB HL or greater hearing loss inclusion criterion as the target trials, using the multiply imputed audiometry data in the full sample
2. We used skin cancer physical exam in the year preceding time zero as a negative treatment control. We expect that the causal effect of this exam on the follow-up biomarkers is null and so estimates that differ substantially from null may indicate residual confounding (e.g., related to healthcare utilisation).
3. If some biomarker outcome data were missing because of unmeasured neurocognitive decline leading to loss to follow-up, this could result in bias. To account for this, we conducted two additional analyes using a modified MI approach, known as NARFCS, wherein shift constants of 0.05 and 0.25 standard deviations (-0.05 and -0.25 in the case of Aβ42/Aβ40) were added to each imputed outcome value.^30,31^
4. We used multivariate adaptive regression splines (with interactions of degree 2) to fit the treatment and outcomes models in TMLE, a more flexible and data-adaptive approach than the primary GLMs.^32^

## RESULTS

A flow diagram describing participant selection is presented in **eFigure 1**. Across imputed datasets, the median (Q1, Q3) sample size of eligible participants for each target trial emulation was 2842 (2808, 2872). A median of 735 (711, 756) participants received a new HA prescription by the start of follow-up. Characteristics of the sample are described in **Table 2**. The mean age was 75 years and 48% were female. The median interval between time zero and the follow-up blood draw was 7.3 years (6.5, 8.1). The follow-up blood draw was precluded by death for a median of 89 participants (12%) who received a new HA prescription and for 209 participants (10%) who did not. The distributions of the biomarker outcomes are displayed in **eFigure 2**.

**Table 2.** Sample characteristics*.

| *Characteristic* | *Summary<sup>1</sup>* |  |
| --- | --- | --- |
|  | *New HA prescription* | *No new HA prescription* |
| N<sup>2</sup> | 735 (711, 756) | 2107 (2078, 2137) |
| Age | 75 (72, 78) | 74 (71, 76) |
| Woman | 360 (49%) | 1014 (48%) |
| White race | 720 (98%) | 2087 (99%) |
| Education |  |  |
| < 9 years | 124 (17%) | 282 (13%) |
| 9-11 years | 226 (31%) | 657 (31%) |
| 12 years | 86 (12%) | 238 (11%) |
| 13-15 years | 108 (15%) | 328 (16%) |
| 16 years | 59 (8%) | 179 (9%) |
| 17-21 years | 130 (18%) | 422 (20%) |
| 3MS Overall score | 94 (91, 97) | 94 (92, 97) |
| BMI, median (IQR) | 28 (25, 30) | 28 (25, 30) |
| History of diabetes | 62 (9%) | 144 (7%) |
| Antihypertensive medication use | 25 (3%) | 61 (3%) |
| Smoking status |  |  |
| Never | 409 (56%) | 1142 (54%) |
| Former | 306 (42%) | 914 (43%) |
| Current | 18 (2%) | 51 (2%) |
| 4-frequency PTA in better ear (dB HL) | 33 (26, 40) | 27 (20, 33) |
| Systolic blood pressure | 139 (128, 151) | 139 (127, 150) |
| APOE ε4 positivity | 179 (24%) | 549 (26%) |
| *Pre-treatment biomarkers* |  |  |
| Risk score (%)<sup>3</sup> | 8 (3, 10) | 8 (3, 10) |
| pTau181 (pg/mL) | 37 (23, 44) | 37 (24, 44) |
| Aβ42/Aβ40 x 1000 | 62 (54, 69) | 62 (54, 69) |
| GFAP (pg/mL) | 133 (89, 161) | 131 (85, 160) |
| NFL (pg/mL) | 22 (15, 25) | 21 (15, 25) |

\* Measured at recruitment into ASPREE/ALSOP study (three years preceding time zero of follow-up). Summary statistics are averaged over the imputed datasets. <sup>1</sup> Mean (Q1, Q3) for numeric variables; n (%) for categorical variables. <sup>2</sup> Median (Q1, Q3) across imputed datasets. <sup>3</sup> Dementia risk score estimated based on plasma biomarkers in out of sample data. PTA: pure tone average. 3MS: Modified Mini-Mental State examination.

### Estimated effects on mean difference scale

The estimated observational analogues of the ITT effects in survivors on the mean difference scale are displayed in **Table 3**. All estimated mean differences were close to null. For example, in the emulation of the first target trial, the estimated mean concentration of pTau181 was 1.8 pg/mL (95% CI: -0.6, 4.1) greater under HA prescription than under no HA prescription. For the emulation of the second target trial, the estimated mean concentration of pTau181 was 2.3 pg/mL (-1.0, 5.5) greater under initiation of always using HAs compared to never using HAs.

**Table 3.**
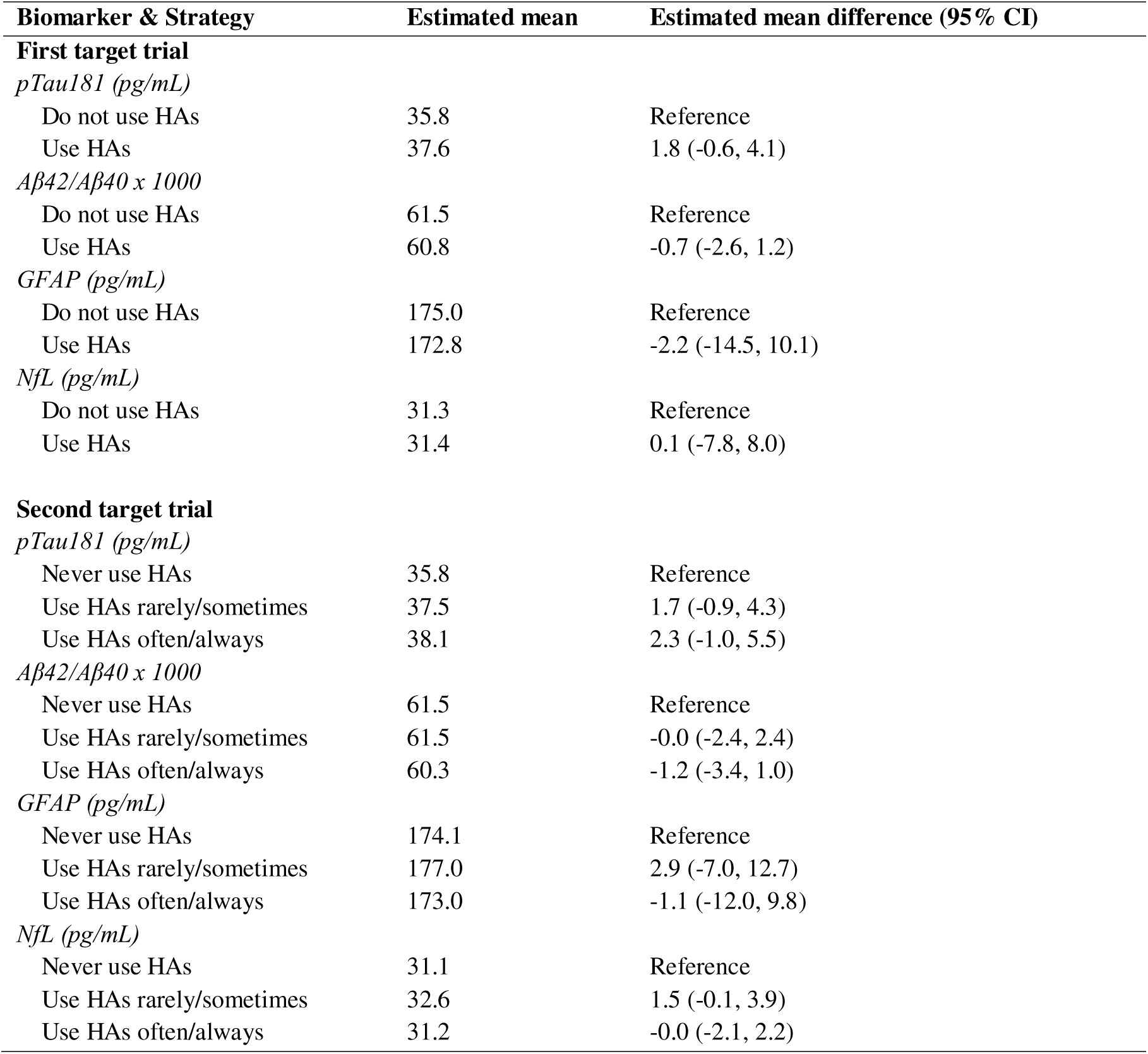
Estimated observational analogues of intention-to-treat mean biomarker concentrations in survivors under each treatment strategy.

### Estimated effects on quantile difference scale

The estimated observational analogues of the ITT effects in survivors on the quantile difference scale are displayed in **Table 4**. All estimated quantile differences were close to null. For example, in the emulation of the first target trial, the estimated 90^th^ percentile of pTau181 was 3.3 pg/mL (-2.3, 8.9) greater under HA prescription than under no prescription.

**Table 4.** Estimated observational analogues of intention-to-treat biomarker concentration percentiles in survivors under each treatment strategy.

| **Biomarker & Strategy** | **Estimated 90<sup>th</sup> Percentile (10<sup>th</sup> percentile for Aβ42/Aβ40)** | **Quantile Difference (95% CI)** |
| --- | --- | --- |
| **First target trial** |  |  |
| *pTau181 (pg/mL)* |  |  |
| Do not use HAs | 55.2 | Reference |
| Use HAs | 58.5 | 3.3 (-2.3, 8.9) |
| *Aβ42/Aβ40 × 1000* |  |  |
| Do not use HAs | 43.8 | Reference |
| Use HAs | 40.5 | -3.3 (-7.1, 0.5) |
| *GFAP (pg/mL)* |  |  |
| Do not use HAs | 279.2 | Reference |
| Use HAs | 274.5 | -4.7 (-29.1, 19.7) |
| *NfL (pg/mL)* |  |  |
| Do not use HAs | 49.2 | Reference |
| Use HAs | 50.7 | 1.5 (-4.2, 7.3) |
| **Second target trial** |  |  |
| *pTau181 (pg/mL)* |  |  |
| Never use HAs | 55.4 | Reference |
| Use HAs rarely/sometimes | 57.2 | 1.9 (-4.3, 8.0) |
| Use HAs often/always | 59.4 | 4.1 (-2.5, 10.7) |
| *Aβ42/Aβ40 × 1000* |  |  |
| Never use HAs | 43.8 | Reference |
| Use HAs rarely/sometimes | 43.7 | -0.2 (-4.1, 3.7) |
| Use HAs often/always | 39.7 | -4.2 (-8.1, -0.3) |
| *GFAP (pg/mL)* |  |  |
| Never use HAs | 279.1 | Reference |
| Use HAs rarely/sometimes | 278.3 | -0.8 (-25.6, 24.1) |
| Use HAs often/always | 271.4 | -7.7 (-30.7, 15.3) |
| *NfL (pg/mL)* |  |  |
| Never use HAs | 49.0 | Reference |
| Use HAs rarely/sometimes | 51.1 | 2.1 (-5.9, 10.1) |
| Use HAs often/always | 50.7 | 1.7 (-4.9, 8.4) |

### Effect modification

The estimated effects of HA prescription did not substantially differ across the levels of the baseline effect modifiers. For example, in those with relatively poor baseline cognition (3MS overall score of 85), the estimated mean concentration of pTau181 was 0.9 pg/mL greater under HA prescription than under no HA prescription. In those with good baseline cognition (3MS overall score of 100), the estimated mean difference was 1.6 pg/mL (**Figure 2**). Findings were similar for the other biomarker outcomes (**eFigures 3-5)**.

**Figure 2.**
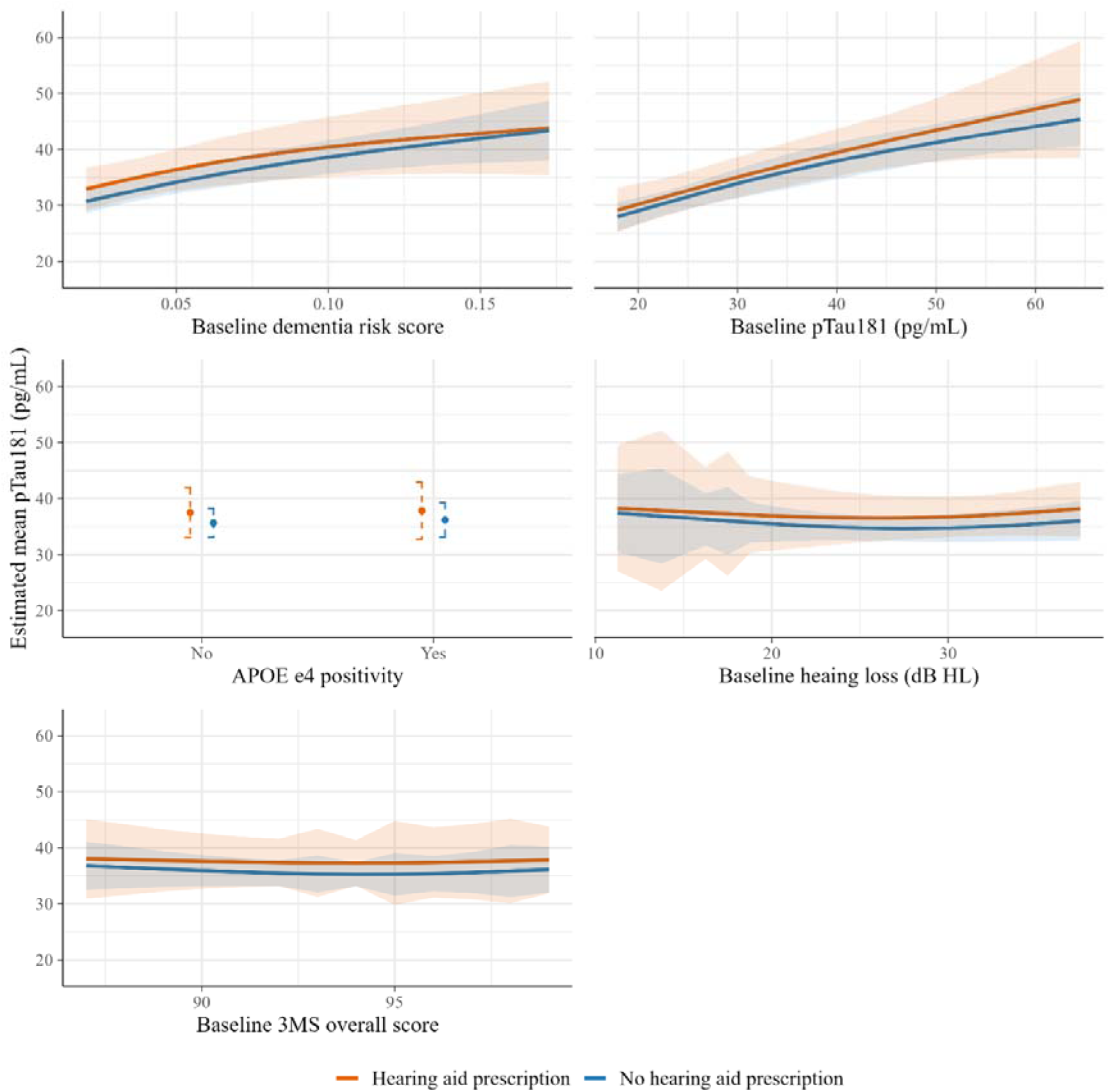
Estimated observational analogues of intention-to-treat mean pTau181 concentrations in survivors under each strategy, by effect modifiers For continuous effect modifiers, the x axis ranges from the 2.5^th^ to the 97.5^th^ percentiles of the effect modifier. Ribbons are bootstrap 95% confidence intervals.

### Sensitivity analyses

Estimated effects of the skin cancer physical exam negative treatment control were close to null for all biomarker outcomes (**eTable 4**). In the sample with hearing loss estimated as 30dB or greater (based on the multiply imputed audiometry data), the results were similar to the main analysis (**eTable 5**). Applying delta adjustment to imputed values for outcomes missing due to loss-to-follow-up also did not substantively alter results (**eTable 6**). Finally, results were similar when using multivariate adaptive regression splines to fit the treatment and outcome models (**eTable 7**).

## Discussion

Our observational analysis found that HA prescription and the initiation of different frequencies of HA use had minimal association with plasma biomarkers of ADRD measured after approximately 7 years of follow-up. Counter to our supposition that treatment effects may be greater among those with elevated pre-treatment ADRD risk, we found that effect estimates tended to differ little by baseline cognition or biomarker concentrations. Finally, we found that the estimated effects of HA prescription and frequency of HA use on the 90^th^ (10^th^ for Aβ42/Aβ40) percentiles of the biomarker distributions were similarly minimal.

There are several plausible explanations for our findings. A first is that there is truly no effect of treating hearing loss with HAs on the aspects of ADRD pathology that are reflected in the studied biomarkers. Instead, the putative benefits of HAs for delaying or preventing dementia may work through other mechanisms, such as through reducing social isolation and depression (both establishment risk factors for dementia), enhancing cognitive reserve through cognitive engagement/stimulation, or via neurological pathways not captured by the biomarkers studied here (e.g., neuroinflammatory).^3^ A second explanation is that, though there could be a real effect of HAs on ADRD pathology, it may be small and/or only detectable with high and sustained HA adherence (which may not reflect the routine clinical setting). Finally, the effect of treating hearing loss with HAs in this older (aged ≥70 years) population may differ from the effect of initiating the treatment at a younger age. Indeed, midlife appears to be a particularly crucial period and interventions initiated at this time may have the greatest benefit.^1^

Though an increasing number of studies are considering the use of plasma biomarkers as dementia surrogate endpoints,^33^ we are not aware of previous studies that have investigated the effect of HAs on the concentration of ADRD biomarkers. Several studies have investigated the association between hearing loss itself and biomarkers of ADRD. Though the overall picture is ambiguous, these tend to suggest that the cerebrospinal fluid (CSF) concentrations of phosphorylated and total tau are modestly elevated in those with hearing loss, but that there is little association between hearing loss and levels of CSF or PET Aβ.^34–36^

Strengths of the study include the long duration of follow-up between HA prescription and biomarker measurement and the use of a study design approach that helps to avoid common biases (e.g., those afflicting prevalent user designs).^31^ Limitations include the use of self-reported hearing problems to emulate ≥30dB HL hearing loss in the primary analysis, an imperfect surrogate.^32^ The hearing loss level for approximately half of the sample was below this 30dB HL level, though most participants had some degree of loss. We also had no information on participants with contraindications for HAs, though we expect this proportion would be small. Secondly, inclusion in the observational sample required that participants met the eligibility criteria for the ASPREE trial, including the absence of cardiovascular disease. The sample data would therefore differ from the target population in its physical health characteristics, a source of bias if these physical health factors were also effect modifiers. Thirdly, the timing of confounder measurement was not perfectly aligned with the time of HA prescription. Residual confounding could result if confounders changed substantially between the time of their measurement at ASPREE/ALSOP baseline and the time of HA prescription (up to 3 years later).^37^

A fourth limitation is the missing data, especially in audiometric hearing loss and the biomarker outcomes. Though we used multiple imputation to handle this missing data, the validity of our inferences is thus sensitive to violations of missing data assumptions. Finally, as we did not have longitudinal data on HA adherence, we were restricted to estimating observational analogues of the ITT effect. It would have been additionally valuable to estimate the effect of sustained HA adherence (i.e., the per-protocol effect), especially given that ITT estimates may not generalise well to other populations/settings with different degrees of adherence.^38^

In conclusion, our study found minimal effect of HA prescription and use on ADRD biomarkers after a 7-year follow-up. Future long-term randomised trials or observational studies could examine the impact of sustained HA adherence and consider other markers of ADRD pathology.

## Data availability

Data from the ASPREE study and associated sub-studies is available to researchers who receive approval for an expression of interest in the ASPREE access management system (https://ams.aspree.org/public/request-data/access-aspree-data/).

## Supporting information

TARGET checklist

## Data Availability

Data from the ASPREE study and associated sub-studies is available to researchers who receive approval for an expression of interest in the ASPREE access management system.

https://ams.aspree.org/public/request-data/access-aspree-data/

## Acknowledgements

The authors are grateful to the ASPREE study participants and the ASPREE study staff for their essential contributions.

## Study funding

This research is supported by funding from the National Institute on Aging (grant R01AG079397 to AM, MM and JR029824). The ASPREE project, comprising two components of ASPREE and ASPREE-XT, was led by Monash University in Australia and the Berman Centre for Outcomes and Clinical Research in the USA. Funding for the ASPREE project was provided by Australian and US governments; the Australian National Health and Medical Research Council (NHMRC) (grants 334047 and 1127060); the National Institute on Aging; the National Cancer Institute at the US National Institutes of Health (grants U01AG029824 and U19AG062682); Monash University (Australia) and the Victorian Cancer Agency (Australia). ALSOP received funding support from Monash University, ANZ Trustees, the Wicking Trust and the Mason Foundation. LC is supported an Australian Government Research Training Program Scholarship and Monash Graduate Excellence Scholarship. MMB was supported by an Australian NHMRC Investigator Grant (ID 2009572). MP received an Australian National Health NHMRC Investigator Grant Fellowship (GTN2009264). GR is supported by the Graeme Clark Chair in Audiology and Speech Science. JR is funded by Leadership 1 Investigator Grant 2016438 from the NHMRC.

## Competing interests

Dr. RJ Shah reports being the site principal investigator or sub-investigator for Alzheimer’s disease clinical trials for which his institution (Rush University Medical Center) is compensated [Athira Pharma, Inc., Annovis Bio, Inc., Edgewater NEXT, Eisai, Inc., and Genentech, Inc.]. Dr. Shah also reports a compensated scientific advisory board agreement with Lundbeck, Inc., and a non-compensated consultant agreement with Novo Nordisk regarding diagnosis and management of dementia. Dr T T.-J Chong reports receiving honoraria for lectures from Roche. None of these are directly related to the work in this manuscript. All other authors report no disclosures.

## Notes

### Clinical Protocols

https://osf.io/9faew/overview

### Author Declarations

ASPREE and ASPREE-XT were approved by the ethics review board at each participating institution. All datasets were fully de-identified prior to use for this study.

