## Appendix for "The Effect of Treating Hearing Loss with Hearing Aids on Plasma Biomarkers of Alzheimer’s Disease and Related Dementias"

### eMethods: Identifying assumptions

The primary causal estimand, for the first emulated target trial, is the observational analogue of the intention to treat effect in survivors on the mean difference scale. I.e., the effect of hearing aid (HA) prescription versus no HA prescription. In potential outcomes notation, we define this as $E\left[ Y^{a=1,m_{Y}=0} \right]-E\left[ Y^{a=0,m_{Y}=0} \right]$, with $A$ representing HA prescription, $M_{Y}$ representing missing outcome data, and *S* representing survival. That is, the causal estimand is the mean difference in the concentration of a given biomarker in survivors under HA prescription and no HA prescription and the absence of missing outcome data. This section describes the assumptions required to identify this causal estimand.

#### Missingness directed acyclic graph

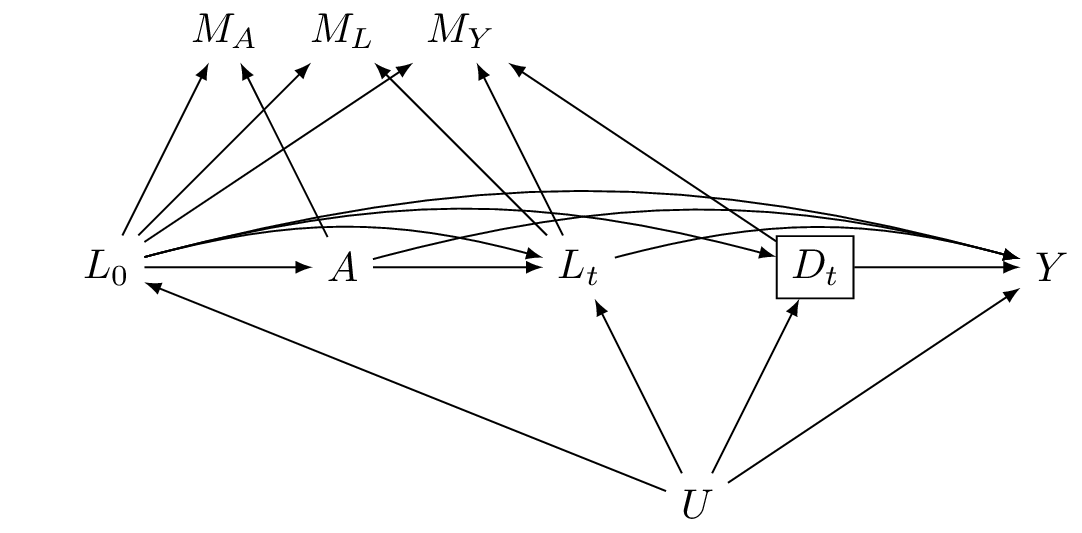

The above figure displays a missingness directed acyclic graph (m-DAG)^1^ for a given biomarker outcome *Y*. *L_0_* is a vector of baseline confounders, *A* is treatment, $L_{t}$ is a vector of covariates at time *t*, $D_{t}$ is death by time *t*, *U* is a vector of unmeasured variables, and $M_{L}$, *M_A_*, and *M_Y_* are indicators for missingness in the covariates, treatment, and outcome, respectively. The rectangle around $D_{t}$ represents conditioning (by restriction to survivors; *D* = 0).

The identifiability of our causal estimand requires the standard causal assumptions: conditional exchangeability, positivity, and consistency, as well as assumptions concerning measurement error. We discuss each of these assumptions in detail below. As missing data is present, however, additional assumptions concerning the structure of this missing data are required for this estimand to be “recoverable”. To examine these missing data assumptions, we make reference to recent theoretical and empirical recoverability results for a set of “canonical” m-DAGs.^1,2^ A key finding of this work is that the average causal effect will not generally be recoverable when any variable is a cause of its own missingness.^2^ For our m-DAG above, this is true for the covariates $L$ and for the exposure $A$. Thus, our causal estimand is non-recoverable. Nevertheless, we believe that our estimand can be estimated with little bias in practice for two main reasons: First, we expect the paths connecting the covariates and treatment with their own missingness indicators to be weak (e.g., an individual’s hearing function, for example, is not a strong determinant of their decision to respond to survey items concerning their hearing function). Second, even for theoretically non-recoverable estimands, empirical findings indicate that appropriate multiple imputation approaches can yield approximately unbiased causal effect estimates, so long as the outcome is not a cause of its own missingness (or that there are no common causes of the outcome and its missingness).^2^ This latter condition holds for our m-DAG as we have that $Y\perp M_{Y}|L_{0},{A,L}_{t},D=0$. Note that this conditional independence would not hold without including the longitudinal auxiliary variables $L_{t}$. In the next paragraphs, we discuss the other assumptions required for identification.

*Conditional exchangeability*

Conditional exchangeability refers to the independence of potential outcomes from the treatment received, given the adjustment variables. For HA treatment, we assume that $Y^{a,m_{Y}=0}\perp A|L_{0},D=0$ for all *a*, i.e., that conditional exchangeability holds given the baseline confounders and survival until biomarker measurement. This assumption is represented in the m-DAG by the absence of unblocked backdoor paths from the outcome to the HA treatment variable after conditioning on these two nodes. Note that, in the m-DAG, there are no direct or indirect paths from treatment to death. Consequently, there are no unblocked backdoor paths created by restricting the analysis to those who survive until outcome measurement. We previously found minimal association between HA use and overall mortality over 7 years and thus believe this assumption is plausible.^4^

#### Positivity

Positivity holds when all participant subgroups within the target population have some positive probability of receiving each treatment level. For this study, we require positivity for missingness and for HA treatment. For missingness, we assume that for all $\left( l_{0},a,l_{t} \right)$ such that $f_{L_{0},A,L_{t},D}\left( l_{0},a,l_{t},0 \right)\neq0$, $\Pr\left[ M=0 | L_{0}=l_{0},A=a,L_{t}=l_{t},D=0 \right]$ > 0 in the target population. I.e., that there are some surviving individuals with non-missing data for every subgroup defined by values of the treatment, baseline covariates, and the auxiliary variables. For HA treatment, we assume that if $f_{L_{0},D}\left( l_{0},0 \right)\neq0$ then $\Pr\left[ A=a | L_{0}=l_{0}, D=0 \right]$ > 0 for all *l_0_* and all *a* among survivors.

#### Consistency

Consistency refers to the potential outcomes under an intervention being equal to the observed outcomes for those who received that intervention*.* I.e., that $Y^{a,m_{Y}=0}=Y$ for those with *A* = *a* and $M_{Y}=0$. In our setting, there are plausibly multiple versions of HA prescription (e.g., behind-the-ear vs in-the-ear HAs) that could result in different biomarker outcomes. Nevertheless, under assumptions that we expect to hold here (i.e., the absence of unmeasured common causes of HA prescription and the version of HA prescription received), our estimand remains estimable and has a natural interpretation: the average effect of HA prescription on the outcome with the versions of HA prescription drawn from the distribution of versions of HA prescription in the target population.^3^

#### Measurement error

**Covariates**: We assume that some of the baseline covariates, like the baseline biomarker concentrations, are measured with error.

**Exposures**: We assume that: i) HA prescription is measured without error and ii) the frequency of HA use is measured with error (as self-report tends to systematically overestimate actual HA use frequency^4^)

**Outcomes**: As the follow-up biomarkers are measured from plasma, they provide only a proxy measure of their CNS concentration. We assume that the measurement error in these biomarkers does not differ by (i.e., is non-differential with respect to) HA prescription and use frequency. Furthermore, we assume that the measurement error in the biomarkers is independent from the measurement error in the HA use frequency exposure.

**Impact of measurement error:** Under these assumptions, measurement error in the confounders will generally introduce bias (due to residual confounding).^5^ Otherwise, the measurement error in the outcome we would not expect to introduce meaningful bias, as independent and non-differential error in a continuous outcome does not bias mean differences.^5^ Measurement error in the HA use frequency variable is likely to introduce bias towards the null as higher use categories (e.g., “always use HAs”) will be constituted by a mix of those with truly high HA use frequency and those with lower use frequencies.^5^

### eMethods: Constructing the biomarker-based dementia risk score

To create the all-cause dementia risk score based on the pre-treatment biomarkers, we used the following algorithm:

1) We created an “external” dataset that contained ASPREE study participants that did not meet the eligibility criteria for this study and who had baseline biomarker data available (n = 8,329 individuals, 785 deaths, and 615 dementia cases).
2) In the external dataset, we create a person-period (discrete time survival) dataset that included, for each participant, as many rows of data as they had 2-year follow-up intervals, ending either at dementia, death, loss to follow-up, or after 10 years
3) In the external dataset, we fitted SuperLearner^15^ models for the discrete time of hazard of dementia and of death, including time and the biomarkers as predictors
4) In the main analysis dataset, we computed the estimated 10-year cumulative incidence of dementia (the risk score) as a function of the hazards from the fitted models in (3)

The use of external data for fitting the models for the risk score prevents bias in the use of the risk score related to overfitting.^6^ Learners included in the SuperLearner for estimating the discrete time hazards were GLMs, generalised additive models, multivariate adaptive regression splines, Bayesian adaptive regression trees, and gradient boosting. All algorithms used default hyperparameters of the SuperLearner R package, except for gradient boosting (minimum size of leaf node set to 25 and maximum tree depth set to 3). 10-fold cross-validation was used to estimate the ensemble weights.^7^ Random forests were pre-specified for inclusion but were removed due to computational limitations.

### eTable 1: Prespecified baseline confounders and model specification

| **Confounder** | **Available ASPREE or ALSOP measures** | **Functional form** | **Product terms in outcome models** |
| --- | --- | --- | --- |
| Time from baseline* | Time between start of follow-up and biomarker measurement | RCS(3) | Treatment |
| Biomarkers of neurodegeneration | pTau-181, NfL, GFAP, Aβ42 / Aβ40, risk score | RCS(3) | Treatment, key covariates^1^, other baseline biomarkers, time from baseline |
| Age | Age | RCS(3) | Treatment, key covariates^1^, baseline biomarkers |
| Gender | Gender | Binary | Treatment, key covariates^1^, baseline biomarkers |
| BMI | BMI | RCS(3) | - |
| Education | Years of education | RCS(3) | Treatment, key covariates^1^, baseline biomarkers |
| Race | Race | Categorical | - |
| Socioeconomic status | Postcode based metric of socioeconomic status (Index of Relative Socio-economic Advantage and Disadvantage) | RCS(3) | - |
|  | Income (4-point scale ranging from <20,000 AUD to >100,000 AUD) | Quadratic | - |
| Social isolation | Sum score of social engagement questionnaire items | RCS(3) | - |
| Physical activity | Physical activity (self-rated; 4-point scale representing physical activity performed in a typical week, ranging from 0 [rarely/never perform physical activity] to 3 [regular vigorous physical activity]) | Quadratic | - |
| Sleep duration | Hours of nightly sleep (self-rated; 6-point scale ranging from <4 hrs to >12hrs) | Quadratic |  |
| Smoking | Smoking status (“current”, “former”, “never”) | Categorical | - |
| Alcohol intake | Typical alcoholic drinks/week | RCS(3) | - |
| Hearing function | Hearing deterioration over last 5 years (self-rated) | Binary | - |
|  | Tinnitus (self-rated; 5-point scale from “never” to “always”) | Quadratic |  |
|  | Difficulty hearing in quiet room (self-rated; 4-point scale from “not at all” to “a lot”) | Quadratic | - |
|  | Difficulty hearing in crowded room (self-rated; 4-point scale from “not at all” to “a lot”) | Quadratic | Treatment |
|  | Pure tone average (PTA) of air conduction thresholds at 0.5, 1, 2, and 4 kHz in the better ear | RCS(3) | Treatment, key covariates^1^, baseline biomarkers |
| Physical health | Physical component score of Short Form-12 | RCS(3) | - |
| Mental health | Mental component score of Short Form-12 | RCS(3) | - |
| Depression | Center for Epidemiological Studies – Depression total score | RCS(3) | - |
| Polypharmacy | Total reported medications | RCS(3) | - |
| History of cancer | Self-reported history of cancer | Binary | - |
| History of diabetes | Self-reported history of diabetes | Binary | - |
| Systolic blood pressure | Systolic blood pressure | RCS(3) | - |
| Cognitive function | 3MS overall score | RCS(3) | Treatment, key covariates^1^, baseline biomarkers |
|  | HVLT-R delayed recall | RCS(3) | Treatment |
| APOE-e4 genotype | APOE-e4 genotype | Linear | Treatment, key covariates^1^, baseline biomarkers |
| Frailty | Deficit-Accumulation Frailty Index | RCS(3) | Treatment, key covariates^1^, baseline biomarkers |
| Visual impairment | 6-point self-rated eyesight scale, with responses ranging from “excellent” to “completely blind” | Quadratic | - |
| Chronic kidney disease | Chronic kidney disease |  | - |
| eGFR | eGFR | RCS(3) | - |
| Liver function | First two principal components of available liver function test results (e.g., AST, ALT, ALP, Bilirubin, GGT) | RCS(3) | - |

Confounders are measured at recruitment into ASPREE or ALSOP studies. * Outcome model only. RCS(3) = restricted cubic spline with knots at the 10^th^, 50^th^, and 90^th^ percentiles. ^1^ Age, gender, education, 3MS overall score, APOE e4 genotype, frailty. Product terms were pre-specified for inclusion in treatment models but were excluded due to model non-convergence in some bootstrap samples.

### eTable 2: Summary of missing data

| **Variable** | **Missing (%)** |
| --- | --- |
| ***Eligibility*** |  |
| Prevalent hearing aid prescription at cohort entry | 4% |
| Self-reported hearing impairment | 5% |
| ***Treatment*** |  |
| New hearing aid prescription (ASPREE year 3) | 18% |
| Hearing aid use (ASPREE year 3) | 18% |
| ***Outcomes*** |  |
| Follow-up biomarkers of ADRD* | 56% |
| ***Covariates*** |  |
| Age | 0% |
| Gender | 0% |
| Race | <1% |
| Education | <1% |
| Income | 18% |
| Socioeconomic status (socioeconomic index for areas) | <1% |
| Baseline biomarkers of ADRD | 27% |
| Difficulty hearing in quiet room | 4% |
| Difficulty hearing in crowded room | 5% |
| 4-Frequency pure tone average | 93% |
| Tinnitus | 9% |
| Systolic blood pressure | 0% |
| Body mass index | <1% |
| eGFR | 3% |
| Liver function | 29% |
| History of cancer | <1% |
| History of diabetes | <1% |
| Antihypertensive use | 0% |
| Polypharmacy | 0% |
| Frailty | 0% |
| Visual function | 2% |
| Chronic kidney disease | 8% |
| Smoking status | 0% |
| Alcohol consumption | <1% |
| APOE-e4 | 17% |
| Typical sleep duration | 2% |
| 3MS Overall score | 0% |
| HVLT-R Delayed recall | <1% |
| CES-D Overall score | <1% |
| SF-12 Mental component score | <1% |
| SF-12 Physical component score | <1% |

***** The proportion of missing data for the follow-up biomarkers of ADRD does not include data unavailable due to death before blood draw could occur.

### eTable 3: Pre-specified auxiliary variables and imputation model specification

| **Covariate** | **Available ASPREE or ALSOP measures** | **ASPREE follow-up year** | **Functional form** | **Product terms** |
| --- | --- | --- | --- | --- |
| ***Baseline*** |  |  |  |  |
| Baseline confounders | See eTable 1 | Baseline | Quadratic terms for continuous/ordinal variables, categorical otherwise | All two-way exposure-covariate, exposure-outcome, outcome-covariate, and covariate-covariate product terms for treatment, gender, education, baseline 3MS, baseline HVLT-R delayed recall, baseline 4-frequency PTA, difficulty hearing in crowded room, baseline frailty, apoe-e4, and the baseline and follow-up biomarkers of ADRD |
| ***Outcomes*** |  |  |  |  |
| Follow-up biomarkers of ADRD | pTau-181, NfL, GFAP, Aβ42 / Aβ40 | Year 10 | Quadratic | As above |
| ***Longitudinal auxiliary variables*** | |  |  |  |
| Hearing function | Hearing deterioration over last 5 years | Year 3 | Binary | - |
|  | Tinnitus | Year 3 | Quadratic | - |
|  | Difficulty hearing in quiet room | Year 3 | Quadratic | - |
|  | Difficulty hearing in crowded room | Year 3 | Quadratic | - |
|  | Pure tone average of air conduction thresholds at 0.5, 1, 2, and 4 kHz in better ear | Year 3 | Quadratic | - |
| Depression | Center for Epidemiological Studies – Depression total score | Years 3, 6, 9 | Quadratic | - |
| Frailty | Deficit-Accumulation Frailty Index | Years 3, 6, 9 | Quadratic | Treatment, age, gender, education |
| Cardiovascular disease | Myocardial infarction, heart failure, or stroke | Years 3, 6, 9 | Binary | - |
| Cancer | Any cancer diagnosis | Years 3, 6, 9 | Binary | - |
| Physical health | Short-form 12 physical health component score | Years 3, 6, 9 | Quadratic | - |
| Mental health | Short-form 12 mental health component score | Years 3, 6, 9 | Quadratic | - |
| Polypharmacy | Number of medications | Years 3, 6, 9 | Quadratic | - |
| Cognitive function | 3MS overall score | Years 3, 6, 9 | Quadratic | Treatment, age, gender, education |
|  | HVLT-R delayed recall | Years 3, 6, 9 | Quadratic | - |
| Dementia | Dementia diagnosis | Years 3, 6, 9 | Binary | Treatment, age, gender, education |

### eTable 4. Skin cancer physical exam negative treatment control

| **Biomarker & Strategy** | **Estimated mean** | **Estimated mean difference (95% CI)** |
| --- | --- | --- |
| *pTau-181 (pg/mL)* |  |  |
| No exam | 36.0 | Reference |
| Exam | 36.6 | 0.6 (-0.6, 1.9) |
| *Aβ42/Aβ40 x 1000* |  |  |
| No exam | 61.3 | Reference |
| Exam | 61.3 | 0.0 (-1.2, 1.1) |
| *GFAP (pg/mL)* |  |  |
| No exam | 173.7 | Reference |
| Exam | 173.5 | -0.2 (-4.8, 4.4) |
| *NfL (pg/mL)* |  |  |
| No exam | 31.1 | Reference |
| Exam | 31.2 | 0.1 (-0.9, 1.2) |

### eTable 5. Estimated observational analogues of intention-to-treat mean biomarker concentrations under each treatment strategy among survivors, using audiometry data to emulate moderate or greater hearing impairment*

| **Biomarker & Strategy** | **Estimated mean** | **Estimated mean difference (95% CI)** |
| --- | --- | --- |
| **First target trial** |  |  |
| *pTau-181 (pg/mL)* |  |  |
| Do not use HAs | 37.1 | Reference |
| Use HAs | 39.3 | 2.1 (-0.8, 5.0) |
| *Aβ42/Aβ40 x 1000* |  |  |
| Do not use HAs | 61.5 | Reference |
| Use HAs | 61.3 | -0.2 (-3.4, 3.0) |
| *GFAP (pg/mL)* |  |  |
| Do not use HAs | 180 | Reference |
| Use HAs | 176 | -4.1 (-16.6, 8.3) |
| *NfL (pg/mL)* |  |  |
| Do not use HAs | 34 | Reference |
| Use HAs | 33 | -0.4 (-3.2, 2.5) |
| **Second target trial** |  |  |
| *pTau-181 (pg/mL)* |  |  |
| Never use HAs | 37.1 | Reference |
| Use HAs rarely/sometimes | 38.7 | 1.6 (-1.3, 4.5) |
| Use HAs often/always | 40.6 | 3.5 (-0.7, 7.7) |
| *Aβ42/Aβ40 x 1000* |  |  |
| Never use HAs | 61.4 | Reference |
| Use HAs rarely/sometimes | 61.1 | -0.3 (-2.7, 2.1) |
| Use HAs often/always | 61.0 | -0.5 (-5.4, 4.5) |
| *GFAP (pg/mL)* |  |  |
| Never use HAs | 179.4 | Reference |
| Use HAs rarely/sometimes | 176.2 | -3.3 (-14.7, 8.2) |
| Use HAs often/always | 179.0 | -0.4 (-13.5, 12.7) |
| *NfL (pg/mL)* |  |  |
| Never use HAs | 33.5 | Reference |
| Use HAs rarely/sometimes | 34.0 | 0.5 (-2.7, 3.7) |
| Use HAs often/always | 33.3 | -0.1 (-3.2, 2.9) |

***** Better-ear 4-frequency [0.5 – 4 kHz] pure tone average [PTA] of ≥ 30 dBHL. Missing audiometry data was handled by multiple imputation in the full sample.

### eTable 6. Estimated observational analogues of intention-to-treat mean biomarker concentrations under each treatment strategy among survivors after delta adjustment*

|  | **Delta = 0.05 SDs*** | **Delta = 0.25 SDs*** |
| --- | --- | --- |
| **Biomarker & Strategy** | **Estimated mean difference**  **(95% CI)** | **Estimated mean difference**  **(95% CI)** |
| **First target trial** |  |  |
| *pTau-181 (pg/mL)* |  |  |
| Do not use HAs | Reference | Reference |
| Use HAs | 1.8 (-0.6, 4.1) | 1.8 (-0.6, 4.1) |
| *Aβ42/Aβ40 x 1000* |  |  |
| Do not use HAs | Reference | Reference |
| Use HAs | -0.6 (-2.6, 1.2) | -0.7 (-2.6, 1.2) |
| *GFAP (pg/mL)* |  |  |
| Do not use HAs | Reference | Reference |
| Use HAs | -2.2 (-14.5, 10.1) | -2.2 (-14.5, 10.1) |
| *NfL (pg/mL)* |  |  |
| Do not use HAs | Reference | Reference |
| Use HAs | 0.1 (-7.8, 8.0) | 0.1 (-7.8, 8.0) |
| **Second target trial** |  |  |
| *pTau-181 (pg/mL)* |  |  |
| Never use HAs | Reference | Reference |
| Use HAs rarely/sometimes | 1.7 (-0.9, 4.3) | -1.7 (-0.9, 4.3) |
| Use HAs often/always | 2.3 (-1.0, 5.5) | 2.3 (-1.0, 5.5) |
| *Aβ42/Aβ40 x 1000* |  |  |
| Never use HAs | Reference | Reference |
| Use HAs rarely/sometimes | 0.0 (-2.4, 2.4) | 0.0 (-2.4, 2.4) |
| Use HAs often/always | -1.2 (-3.4, 1.0) | -1.2 (-3.4, 1.0) |
| *GFAP (pg/mL)* |  |  |
| Never use HAs | Reference | Reference |
| Use HAs rarely/sometimes | -2.9 (-7.0, 12.7) | 2.9 (-7.0, 12.7) |
| Use HAs often/always | -1.1 (-12.0, 9.8) | -1.1 (-12.0, 9.8) |
| *NfL (pg/mL)* |  |  |
| Never use HAs | Reference | Reference |
| Use HAs rarely/sometimes | 1.5 (-0.9, 3.9) | 1.5 (-0.9, 3.9) |
| Use HAs often/always | 0.0 (-2.1, 2.2) | 0.0 (-2.1, 2.2) |

*** S**hift constants of 0.05 and 0.25 standard deviations (-0.05 and -0.25 in the case of Aβ42/Aβ40), respectively, were added to each imputed outcome value for participants who were lost to follow-up.

### eTable 7. Estimated observational analogues of intention-to-treat mean biomarker concentrations under each treatment strategy among survivors estimated using multivariate adaptive regression splines

| **Biomarker & Strategy** | **Estimated mean** | **Estimated mean difference (95% CI)** |
| --- | --- | --- |
| **First target trial** |  |  |
| *pTau-181 (pg/mL)* |  |  |
| Do not use HAs | 35.9 | Reference |
| Use HAs | 37.5 | 1.5 (-0.8, 3.8) |
| *Aβ42/Aβ40 x 1000* |  |  |
| Do not use HAs | 61.5 | Reference |
| Use HAs | 60.6 | -0.8 (-3.6, 1.9) |
| *GFAP (pg/mL)* |  |  |
| Do not use HAs | 176.0 | Reference |
| Use HAs | 172.8 | -3.2 (-95.2, 88.8) |
| *NfL (pg/mL)* |  |  |
| Do not use HAs | 31.4 | Reference |
| Use HAs | 31.4 | 0.0 (-2.4, 2.4) |
| **Second target trial** |  |  |
| *pTau-181 (pg/mL)* |  |  |
| Never use HAs | 35.9 | Reference |
| Use HAs rarely/sometimes | 36.8 | 0.9 (-1.3, 3.0) |
| Use HAs often/always | 38.0 | 2.1 (-1.0, 5.2) |
| *Aβ42/Aβ40 x 1000* |  |  |
| Never use HAs | 61.6 | Reference |
| Use HAs rarely/sometimes | 61.0 | -0.6 (-2.4, 1.2) |
| Use HAs often/always | 60.2 | -1.4 (-4.2, 1.5) |
| *GFAP (pg/mL)* |  |  |
| Never use HAs | 174.9 | Reference |
| Use HAs rarely/sometimes | 176.5 | 1.7 (-32.9, 36.2) |
| Use HAs often/always | 172.9 | -1.9 (-14.9, 11.0) |
| *NfL (pg/mL)* |  |  |
| Never use HAs | 31.2 | Reference |
| Use HAs rarely/sometimes | 32.9 | 1.7 (-1.3, 4.7) |
| Use HAs often/always | 31.1 | -0.1 (-2.3, 2.0) |

***** Better-ear 4-frequency [0.5 – 4 kHz] pure tone average [PTA] of ≥ 30 dBHL

### eFigure 1. Flow diagram

Australian participants of ASPREE trial (n=16,703)

Participated in ALSOP cohort (n=14,908)

Completed follow-up blood draw (n=845)*

Completed follow-up blood draw (n=268)*

- Prior treatment for hearing loss (n=4,206)*
- Death/dementia before ALSOP year 3 (n=405)
- No self-reported hearing problems (n=7,455)*

Survived until time of follow-up blood draw (n=1,896)*

No hearing aid prescription reported in ALSOP year 3 (n=2,107)*

Survived until time of follow-up blood draw (n=645)*

New hearing aid prescription reported in ALSOP year 3 (n=735)*

Included (n=2,842)*

* Median across imputed datasets

### eFigure 2. Density of biomarker outcomes by new HA prescription

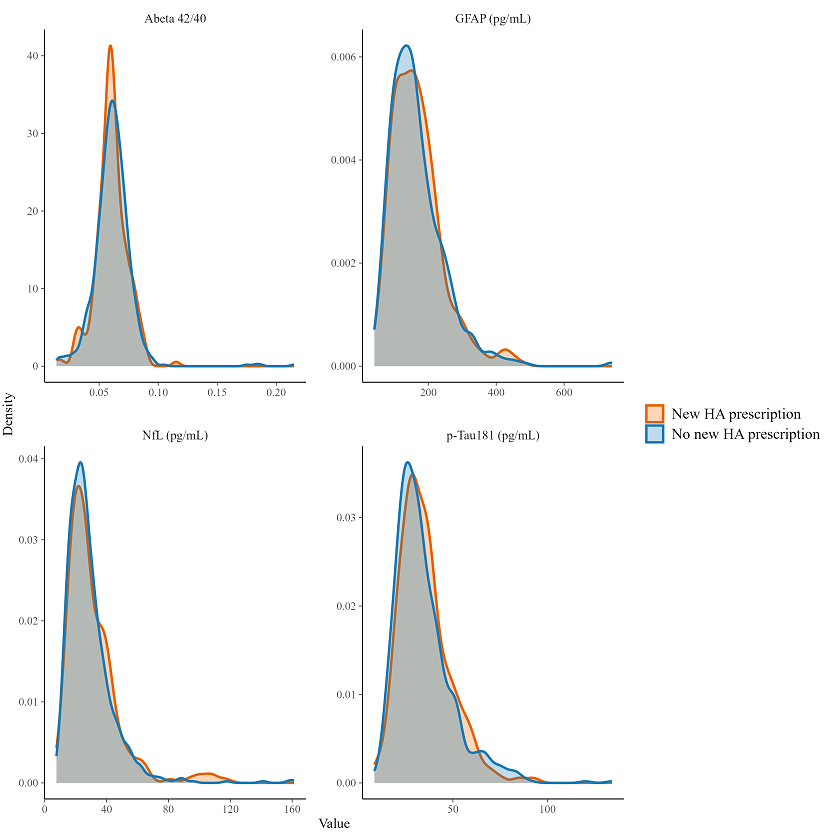

Density plots are obtained from eligible sample with complete exposure and outcome data (n = 999) and are not adjusted for confounding. For visual clarity, outcomes are truncated at 0.1 and 99.9 percentiles.

### eFigure 3. Estimated observational analogues of intention-to-treat mean Aβ42/Aβ40 in survivors under each strategy, by effect modifiers

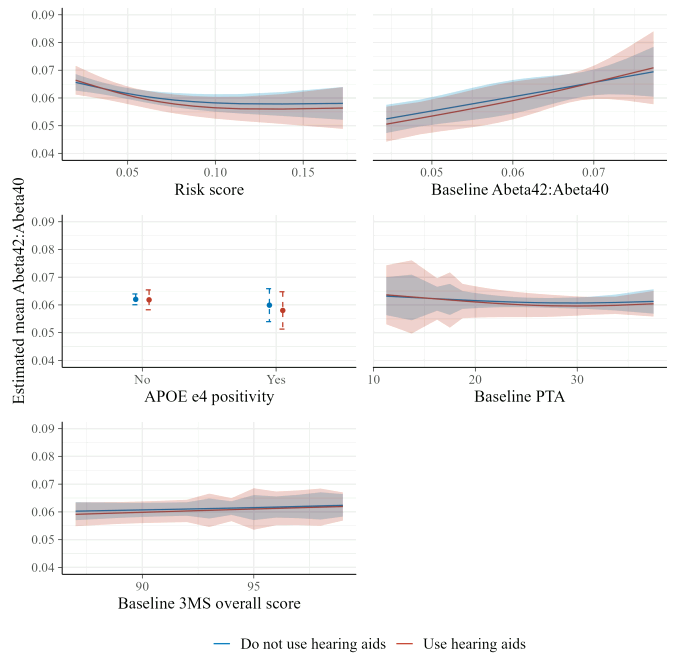

### eFigure 4. Estimated observational analogues of intention-to-treat mean GFAP in survivors under each strategy, by effect modifiers

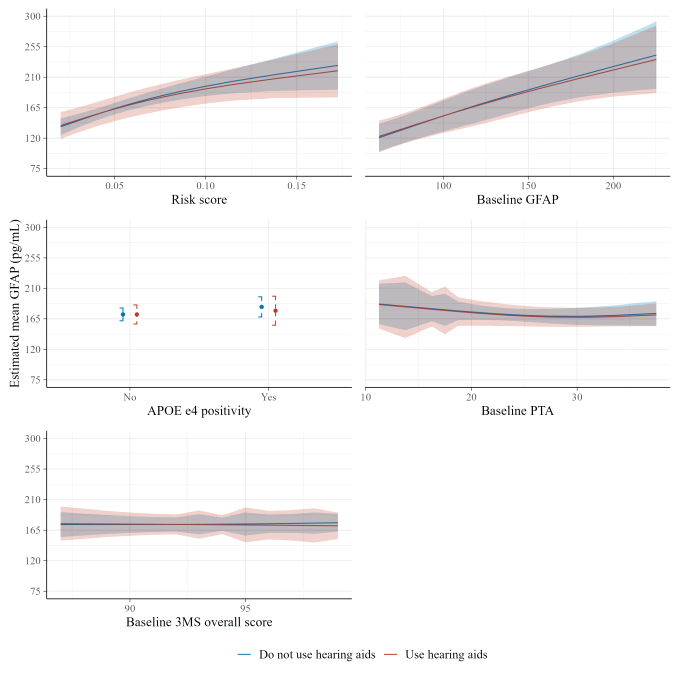

### eFigure 5. Estimated observational analogues of intention-to-treat mean NfL in survivors under each strategy, by effect modifiers

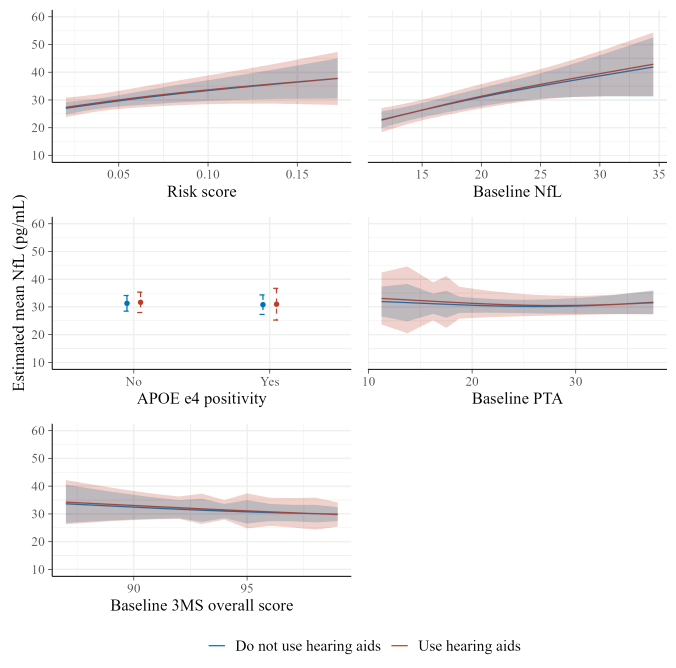
