## Supplementary material for "The Effect of Treating Hearing Loss with Hearing Aids on Plasma Biomarkers of Alzheimer’s Disease and Related Dementias": TARGET checklist

| Item no. | | | | Checklist item | | | | | Location reported | |
| --- | --- | --- | --- | --- | --- | --- | --- | --- | --- | --- |
| Abstract | | | | | | | | | | |
| 1 | a | | | | Identify that the study attempts to emulate a target trial using observational data. State the study objectives and briefly summarize the specified target trial. | | | | 3 | |
|  | b | | | | Report the data sources used for emulation. | | | | 3 | |
|  | c | | | | Summarize key assumptions, statistical methods, findings and conclusions. | | | | 3 | |
| Introduction | | | | | | | | | | |
| 2 | Background | | | | Describe the scientific background of the study and the gap in knowledge. | | | | 5 | |
| 3 | Causal question | | | | Summarize the causal question. | | | | 6 | |
| 4 | Rationale | | | | Describe the rationale for emulating a target trial with the available data. Cite randomized trials informing the design of the target trial if applicable. | | | | 5-6 | |
| **Methods** | | | | | | | | | | |
| 5 | Data sources | | Cite the data sources contributing to the analyses and for each one describe the following: original purpose, type, the geographic locations, setting and time-period. If relevant, describe how the data were linked or pooled. | | | | | | 6 | |
| 6 | Target trial specification  Specify the components of the target trial protocol that would answer the causal question. | | | | | 7 | **Target trial emulation**  Describe how the components of the target trial protocol were emulated with the observational data, including how all variables were measured or ascertained. | | **Location item 6 (specification) reported** | **Location item 7 (emulation) reported** |
|  | Eligibility criteria | | | | |  | Eligibility criteria | | 8 | 8 |
|  | a | Describe the eligibility criteria. | | | |  | a | Describe how the eligibility criteria were operationalized with the data. |  |  |
|  | **Treatment strategies** | | | | |  | **Treatment strategies** | | **8** | **8** |
|  | b | Describe the treatment strategies that would be compared. | | | |  | b | Describe how the treatment strategies were operationalized with the data. |  |  |
|  | **Assignment procedures** | | | | |  | **Assignment procedures** | | **8-9** | **8-9** |
|  | c | Report that eligible individuals would be randomly assigned to treatment strategies and may be aware of their treatment allocation. | | | |  | c | Describe how assignment to treatment strategies was operationalized with the data. |  |  |
|  | **Follow-up** | | | | |  | **Follow-up** | | **9** | **9** |
|  | d | Clarify that follow-up would start at time of assignment to the treatment strategies. Specify when follow-up would end. | | | |  | d | Clarify that follow-up starts at the time individuals were assigned to the treatment strategies. Describe how the end of follow-up was operationalized with the data. |  |  |
|  | **Outcomes** | | | | |  | **Outcomes** | | **9** | **9** |
|  | e | Describe the outcomes. | | | |  | e | Describe how the outcomes were operationalized with the data. |  |  |
|  | Causal contrasts | | | | |  | Causal contrasts | | 9 | 10 |
|  | f | Describe the causal contrasts of interest, including effect measures. | | | |  | f | Describe how the causal contrasts were operationalized with the data, including effect measures. |  |  |
|  | **Identifying assumptions** | | | | |  | **Identifying assumptions** | | **10, S2-S5** | **10, S2-S5** |
|  | g | Describe assumptions that would be made to identify each causal estimand. Describe the variables, if any, related to these assumptions. | | | |  | g.i | For each causal estimand, describe assumptions made to identify it, including assumptions regarding baseline confounding due to lack of randomization. |  |  |
|  |  |  | | | |  | g.ii | Describe how the variables related to these assumptions were operationalized with the data |  | S7-S8, S10-S11 |
|  | **Data analysis plan** | | | | |  | **Data analysis plan** | | - | **10-12** |
|  | h | For each causal estimand, describe the data analysis procedures and any associated statistical modelling assumptions, including approaches for handling missing data. | | | |  | h.i | For each causal estimand, describe the data analysis procedures and any associated statistical modelling assumptions, including approaches for handling missing data. |  |  |
|  |  |  | | | |  | h.ii | For each causal estimand, describe any additional analyses conducted to assess the sensitivity of the results to the choice of operationalizations, assumptions and analysis. |  | 13 |
| **Results** | | | | | | | | | | |
| 8 | Participant selection | | | | Report numbers of individuals assessed for eligibility, eligible, and assigned to each treatment strategy. A flow diagram is strongly recommended. | | | | 13, S16 | |
| 9 | Baseline data | | | | Describe the distribution of characteristics of individuals at baseline, by treatment strategy. | | | | 25 | |
| 10 | Follow-up | | | | Summarize length of follow-up and describe reasons for end of follow-up for each treatment strategy and causal contrast. | | | | 11, 13 | |
| 11 | Missing data | | | | Describe the frequency of missing data in all variables, by treatment strategy when applicable. | | | | 11, S16 | |
| 12 | Outcomes | | | | Describe the frequency or distribution of each outcome, by treatment strategy. | | | | S17 | |
| 13 | Effect estimates | | | | Report the effect estimates for each causal contrast with corresponding measures of precision, including both absolute and relative measures of effect, when applicable. | | | | 14, 26-27, Figure 2 | |
| 14 | Additional analyses | | | | Report results of all analyses to assess the sensitivity of the estimates to choices in operationalizations, assumptions and analysis. | | | | 14-15 | |
| **Discussion** | | | | | | | | | | |
| 15 | Interpretation | | Provide an interpretation of the key findings. | | | | | | 15 | |
| 16 | Limitations | | Discuss the limitations of the study considering differences between the target trial and its emulation and the plausibility of assumptions, including assumptions regarding baseline confounding due to lack of randomization. | | | | | | 16-17 | |
| **Other information** | | | | | | | | | | |
| **17** | Ethics | | | | Provide the institutional research board or ethics committee that approved the study and approval numbers, if relevant. | | | | 7 | |
| **18** | Registration | | | | State whether, when and where the study protocol was registered. | | | | 10 | |
| **19** | Sharing of study materials | | | | Provide information on whether data, analytic code and/or other materials are accessible, and where and how they can be accessed. | | | | 17 | |
| **20** | Funding sources | | | | Provide the sources of funding and detail the role of the funders in the design, conduct and reporting of the study. | | | | 17-18 | |
| **21** | Conflicts of interest | | | | State any conflicts of interest and financial disclosures for all authors. | | | | 2 | |

Citation: Cashin AG, Hansford HJ, Hernán MA, Swanson SA, Lee H, Jones MD, et al. Transparent Reporting of Observational Studies Emulating a Target Trial: The TARGET Statement. JAMA. 2025; DOI: 10.1001/jama.2025.13350
© 2025 Cashin et al. This is an Open Access article distributed under the terms of the Creative Commons Attribution-NoDerivatives License (CC BY-ND 4.0), which permits redistribution, commercial and non-commercial, provided the work is passed along unchanged and in whole, with credit to the original author(s).
